# Aberrant naive CD4+ T Cell differentiation in systemic juvenile idiopathic arthritis is committed to B cell help

**DOI:** 10.1101/2022.02.01.22270100

**Authors:** Julia Kuehn, Susanne Schleifenbaum, Antje Hellige, Carolin Park, Claas Hinze, Helmut Wittkowski, Dirk Holzinger, Dirk Foell, Christoph Kessel

## Abstract

**Objective:** Systemic juvenile idiopathic arthritis (sJIA) features characteristics of autoinflammation and autoimmunity, culminating in chronic arthritis. Previous work indicated decreased IFNγ-expression by T helper (Th) cells in sJIA. Here, we hypothesized this to result from aberrant or incomplete Th cell polarization.

**Methods:** Cells or sera were obtained from healthy controls (HC, n=26) and sJIA patients (n=61). Isolated naïve Th cells were cultured under Th1, Th17, and T follicular or T peripheral helper (Tf/ph) polarizing conditions and were partly co-cultured with allogenic memory B cells. Surface marker, transcription factor, and/or cytokine expression in peripheral or polarized Th cells or sera as well as B cellular plasma blast generation were studied by flow cytometry, multiplexed bead array assays, ELISA and retrospective RNA profiling analyses.

**Results:** SJIA naive Th cell differentiation towards Th1 resulted in low IFNγ and Eomesodermin expression. Instead, developing sJIA Th1 cells revealed elevated IL-21 release that negatively correlated with cellular IFNγ and Eomesodermin expression. Both *In vitro* and *ex vivo*, IL-21 together with PD-1, ICOS and CXCR5 expression indicated naïve sJIA Th cell differentiation to rather polarize towards a Tf/ph phenotype. Retrospective analysis of whole blood RNA sequencing data demonstrated sJIA-specific overexpression of Bcl-6 as respective master transcription factor. Compared to controls, *in vitro* generated sJIA Tf/ph cells promoted enhanced B cellular plasma-blast generation.

**Conclusion:** In sJIA pathogenesis skewing of sJIA naïve Th cell differentiation towards a Tf/ph phenotype may represent an echo of autoimmunity, which could shed light on the mechanisms driving the progression towards chronic destructive arthritis.

## Introduction

Systemic juvenile idiopathic arthritis (sJIA), or Still’s disease, is a unique childhood arthritis entity in that it features characteristics of both autoinflammation and autoimmunity(1). Recent studies indicated that sJIA shows a unique genetic architecture, which manifests in a clinical phenotype distinct from other forms of juvenile arthritis(2). Pathogenetic understanding in sJIA is, however, still limited(3).

At disease onset, the clinical presentation of sJIA is dominated by features prevalent in autoinflammatory fever syndromes. Common symptoms comprise quotidian spiking fever accompanied by an erythematous rash, whereas arthritis may be minimal or even absent. Hepatosplenomegaly, lymphadenopathy, and/or serositis are additional clinical hallmarks that separate sJIA from the other JIA subtypes(4). SJIA can be complicated by macrophage activation syndrome (MAS), a severe hyperinflammatory cytokine storm condition that can result in multi-organ failure with significant mortality(5). If untreated or treatment-refractory, sJIA can progress to chronic destructive arthritis, clinically resembling autoimmunity-driven arthritis(5, 6).

Substantial data underpin the predominance of innate immunity in the early systemic phase of sJIA(3). Therapeutic IL-1 blockade significantly improves disease outcomes(7) and seems particularly effective when initiated as first-line therapy during the systemic disease phase(8–10). Current data suggest, that children with established polyarthritis are less likely to respond to treatment with recombinant IL-1 receptor antagonist (IL-1Ra; anakinra) (11, 12), but may rather benefit from therapeutic IL-6 receptor (IL-6R) blockade (tocilizumab)(13, 14). According to the bi-phasic model of sJIA pathogenesis, innate immunity drives early systemic inflammation but can eventually prime (innately) adaptive immune cells which may then promote chronic destructive arthritis(3, 6); immediate biologic treatment in the early systemic phase has been proposed as “a window of opportunity” to abrogate arthritis evolution(6). A genetic association of sJIA with the major histocompatibility complex (MHC) class II specific allele *HLA-DRB1*11*(15), Th17 cell reprogramming(16) as well as alterations in adaptive immune cell signatures(17) or cells rather bridging innate and adaptive immunity (γδT cells, NK cells)(17, 18) may indeed suggest an autoimmune component to sJIA(6, 19). A growing body of evidence points to a special, supposably dichotomous role for the T cellular cytokine IFNγ and IFNγ-signaling in the sJIA-specific continuum of inflammation. Murine models suggested IFNγ as a key driver of the anemia in MAS(20) and IFNγ-inhibition reverted clinical and laboratory MAS features(21, 22). Similarly, patient data demonstrated that IFNγ-neutralization or inhibition of downstream signaling can control MAS(23–26).

Although a crucial role of IFNγ in MAS pathophysiology is indisputable, its role in active sJIA without MAS is less clear. Analyzing cytokine levels in active sJIA without present or future MAS indicated either no(27–29) or only partial elevation(30, 31) of IFNγ or CXCL9 (as surrogate of IFNγ signaling). Monocytes obtained from active sJIA patients rather pointed to a limited *in vivo* exposure to IFNγ(32, 33). Further, a sJIA mouse model requires absence of IFNγ (IFNγ^-/-^) to induce disease(34, 35). Collectively, these data rather suggested a limited or even opposing role for IFNγ in sJIA development(33); it may even protect from sJIA progression as IFNγ has been shown to inhibit IL-1 signaling in PBMCs(30).

We have previously observed decreased IFNγ expression in CD4+ T cells of both active and inactive sJIA patients(18). In the present study we hypothesized that due to the likely lack of cognate sJIA-associated T cell antigens within an *in vivo* environment of proinflammatory mediators driving T cell polarization into different directions, CD4+ T cells in sJIA may suffer from aberrant or incomplete polarization, which may translate into insufficient IFNγ expression.

## Patients & Methods

### Study individuals

Blood samples and clinical data for all cell experiments were collected from sJIA patients (n=27, active disease (AD), n = 11; inactive disease (ID), n = 16) and pediatric HCs (n = 15) at the University Hospital in Münster, Germany between March 2016 and November 2021. The study was approved by the local ethics committee (2015-670-f-S) and all parents or care givers singed written informed consent. Systemic JIA patients were diagnosed according to the German consensus criteria(36). Active disease (AD) was defined as the presence of arthritis and/or fever and/or rash, while the absence of arthritis, fever, and rash was classified as inactive disease (ID). Patients diagnosed with macrophage activation syndrome (MAS) met the 2016 classification criteria for MAS complicating sJIA(37, 38). Serum samples were collected from sJIA patients (n=34; active disease (AD), n = 25; inactive disease (ID), n = 9) and pediatric HCs (n = 10) in course of a larger study as recently reported by us(39). Collective demographic and clinical characteristics are summarized in **Table 1**.

**Table 1.** Study participants

| Analysis | Disease status | Study subjects, n | Demographics |  |  | Clinical characteristics |  |  | Laboratory parameters |  | Treatment,n |
| --- | --- | --- | --- | --- | --- | --- | --- | --- | --- | --- | --- |
|  |  |  | Age at sampling, median (range) | Sex, n female/male | Median disease duration (range) | No. of active joints (range) | fever, n | rash, n | Median BSR, mm/h (range) | Median CRP, mg/dl (range) |  |
| *in vitro* Th1/Th17 differentiation and *ex vivo* FACS* | HC | 7 | 11 (7 – 16) | 4/3 | NA | NA | NA | NA | NR | NR | NA |
|  | AD | 5 | 8 (6 – 17) | 3/2 | 1 (0.1 – 4) | 3 (0 – 14) | 3 | 2 | 22 (8 – 105) | <0.5 (<0.5 – 6.3) | Toc, 3<br>Sar, 1<br>Pred, 4<br>CsA, 1<br>Tac, 1 |
|  | ID | 9 | 9 (4 – 22) | 6/3 | 6 (0.3 – 11) | 0 | 0 | 0 | 3 (1 – 20) | <0.5 (<0.5 – 1.57) | naïve, 2<br>Ana, 1<br>Cana, 3<br>Toc, 2<br>Ada, 1<br>Pred, 3<br>CsA, 1<br>MTX, 3 |
| *in vitro* Th1/Tfh differentiation and *ex vivo* FACS | HC | 5 | 12 (10 – 22) | 2/3 | NA | NA | NA | NA | NR | NR | NA |
|  | AD | 4 | 12 (7 – 17) | 2/2 | 1.2 (0.2 – 7) | 1 (0 – 8) | 3 | 2 | 13.5 (2 – 55) | <0.5 (<0.5 – 6.0) | naïve, 1<br>Ana, 2<br>Toc, 2<br>Pred, 2<br>CsA, 1<br>MTX, 1 |
|  | ID | 4 | 13.5 (5 – 18) | 2/2 | 1.5 (0.6 – 2) | 0 | 0 | 0 | 9.5 (2 – 12) | <0.5 | naïve, 1<br>Ana, 2<br>Cana, 1<br>Pred, 1<br>CsA, 1<br>Tac, 1 |
| *in vitro* Th1/Tfh | HC | 4 | 23 (22-24) | 2/2 | NA | NA | NA | NA | NR | NR | NA |
| differentiation<br>+<br>memory B cell<br>co-cultures | AD** | 2 | 13 (9.0-19.0) | 2/3 | 2.1 (0.3-4.5) | 0 | 2 | 0 | 59; 19 | >0,5; 0,8 | Ana, 1<br>Pred, 2 |
|  | ID** | 3 |  |  |  | 0 | 0 | 0 | 5 (4-11) | <0.5 | naïve, 1<br>Ana, 1<br>Cana, 1<br>CsA, 1 |
| *ex vivo*<br>cytokines | HC | 10 | 12 (10 – 22) | 2/3 | NA | NA | NA | NA | NR | NR | NA |
|  | AD | 25 | 7 (0.5-18) | 14/11 | 0.65 (0-7) | 1 (0-17) | NR | NR | 80 (28-108) | 8.4 (5.43-76.17) | naïve, 22<br>Ana, 2<br>CsA, 1 |
|  | ID | 9 | 11 (6-18) | 6/3 | 4.5 (0.24-13) | 0 | NR | NR | 5.5 (3-9) | <0.5 | naïve, 7<br>Ana, 2 |

NA not applicable
NR not recorded
Age and disease duration in years
\*The same study samples were used for *ex vivo* flow cytometry analyses of PBMCs and isolation of Th cells for *in vitro* Th cell differentiation experiments and subsequent flow cytometry and cytokine bead assay
\*\*demographics are pooled to allow for full anonymization

### Isolation, (co-)cultures and stimulation of naïve T helper (Th) cells

Naïve Th cells were isolated from PBMCs and stimulated as described in the **supplementary methods section**.

### Flow cytometry, multiplexed bead array assay and ELISA

Flow cytometry of *in vitro* differentiated cells or PBMCs and multiplexed bead array assays or ELISA of sera and cell culture supernatants were performed as detailed in the **supplementary methods section**.

### Statistical analyses

Data analyses were performed using GraphPad Prism software (Version 8.0 for Mac OS X, Graphpad Software, La Jolla, CA, USA) and R studio as detailed in the **supplemental methods section** and figure legends. Corrected *P* < 0.05 was considered statistically significant.

## Results

### Naive CD4+ T cell differentiation in sJIA to Th1 and IFN**γ** production is impaired and skewed toward low Eomesodermin expression

Naive CD4+ T cells were isolated from HCs (n=7) and active (n=5) as well as disease-inactive (n=9) sJIA patients (**Table 1**). Cells were driven toward either Th1 (**Figure 1A, B**) or Th17 polarization (**Figure S2A, B**) by indicated conditions. Subsequent to all T cell polarizations, cells were super-stimulated using PMA/ionomycin and tested for expression of IFNγ, IL-17A, T-bet, RORγt and Eomesodermin by flow cytometry. T cell activation with anti-CD3/CD28 without polarizing cytokines resulted in low IFNγ, IL-17A and Th1/Th17 transcription factor expression (**Figure 1B, C**; **S2B, C**). Yet, IFNγ was tightly associated with T-bet expression (**Figure 1F**). Following Th1 polarization including IL-2 and IL-12 stimulation, IFNγ expression by healthy control naïve T cells was markedly increased over cytokine production by sJIA T cells, particularly from patients with inactive disease (**Figure 1B, D**) and was tightly associated with Eomesodermin rather than T-bet expression (**Figure 1G**). A similar IFNγ expression pattern and IFNγ-Eomesodermin association was observed when including IL-17A (**Figure 1 B, E, H**) or IL-18 (**Figure 1B**, **S1**) into the Th1 polarizing cytokine cocktail. In contrast to IFNγ, both IL-17A and RORγt expression was hardly elevated in all tested Th17 polarizing conditions (**Figure S2B, C**). Compared to healthy controls and inactive disease patients, cells obtained from patients in course of active sJIA tended to express more IL-17A (**Figure S2B, C**).

**Figure 1.**
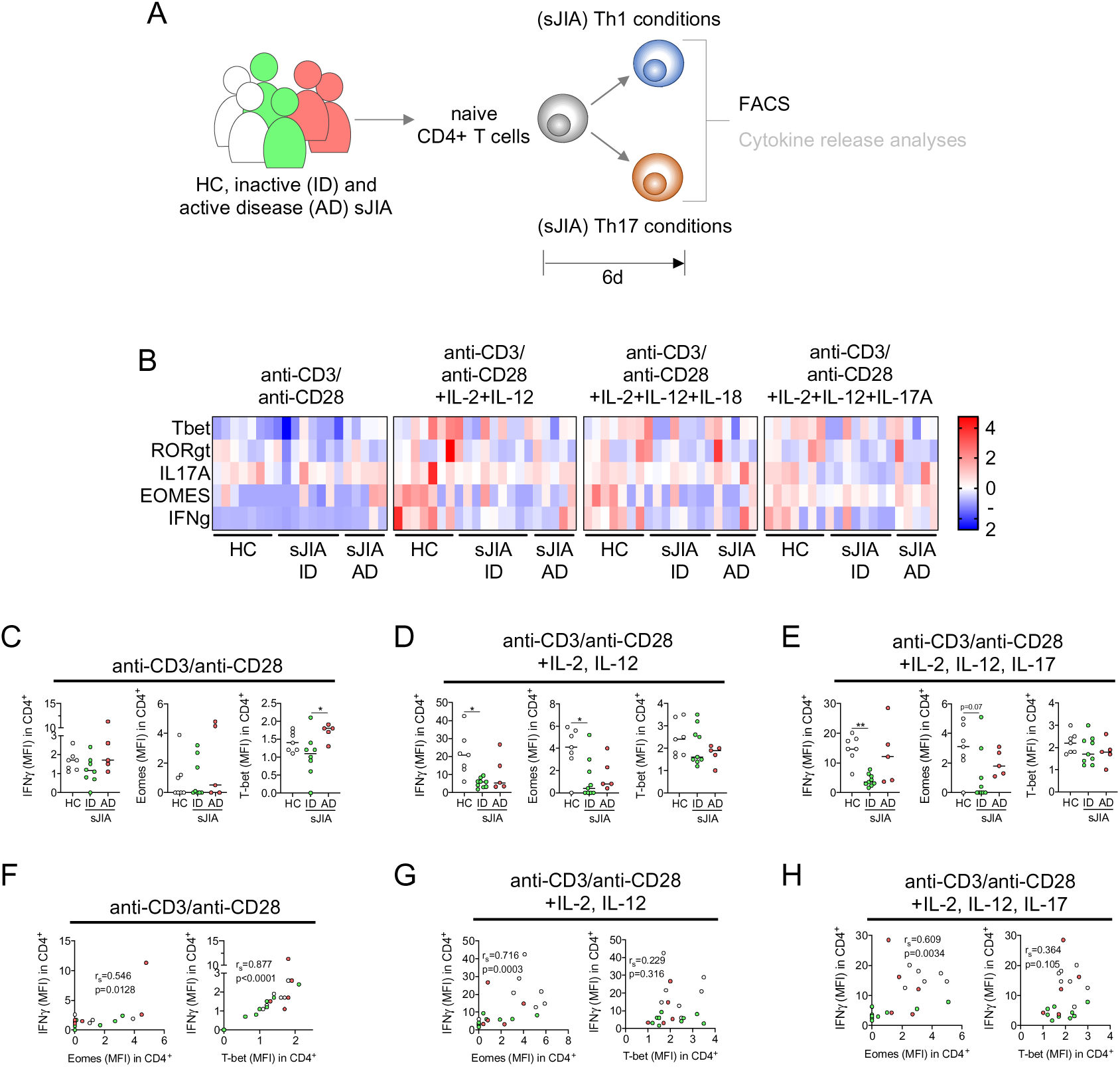
SJIA naïve CD4+ T helper (Th) cell differentiation towards Th1 results in low IFNγ and Eomesodermin expression. **A**, Naive Th cells were isolated from pediatric healthy controls (HC, n=7), active disease (AD, n=5), and inactive disease (ID, n=8) sJIA patients (see table 1) and cultured under T0 (anti-CD3/anti-CD28) and different Th1 and Th17 polarizing conditions. IL-17A and IL-18 were included in Th1 conditions to somewhat mimic a sJIA-cytokine milieu. Following six days of culture and super-stimulation with phorbol 12- myristate 13-acetate/ionomycin (4h), cells were analyzed by flow cytometry. **B**, Heatmap of z-scores indicating T-bet, RORγt, IL-17A, IFNγ, and Eomesodermin (Eomes) expression (geometric mean fluorescence intensity (MFI) normalized to unstimulated samples) in cells arising from the indicated culture conditions and following super-stimulation as described. **C**-**E**, IFNγ (left panels), Eomes (center panels), and T-bet (right panels) expression (MFI), as well as (**F**-**H)** correlation of IFNγ with Eomes (left panels) or T-bet (right panels) expression (MFI) in cells as in **B** is shown. **B**-**H**, Each data point represents a value derived from an individual patient or HC. **C**–**E**, Lines indicate median values, data were analyzed by Kruskal-Wallis test followed by Dunn’s post hoc test; * = *P* < 0.05, ** = *P* <0.01. **F**-**H,** Simple linear regression (OLS) and F-test; r_s_ (r-squared value) and *P*-values are indicated.

Thus, based on cellular expression, we observed only marginal IFNγ and Eomes expression particularly among cells obtained from inactive sJIA patients when driven toward Th1 differentiation.

### Developing sJIA Th1 cells reveal elevated IL-21 release in a negative correlation with IFN**γ** and Eomesodermin expression

Apart from analyzing intracellular cytokine and transcription factor expression, we also quantified IFNγ, IL-17A, IL-21 and IL-22 release if enough cells to prepare respective samples were available (**Figure 2A, B**, **S3A**). Except for one active sJIA patient with future MAS, we observed naïve sJIA T cells stimulated only by CD3/CD28-ligation to release less IFNγ compared to healthy controls (**Figure 2C**, left panel; **S3B**). Conversely, we noted elevated IL-21 production reciprocal to IFNγ release (**Figure 2C**). We further observed, that the phenotype of low IFNγ expression by sJIA T cells was lost with increasing cytokine concentrations in supernatants in course of Th1 differentiation (**Figure 2B, D-E**: left panels), thus contrasting our flow cytometry data at intracellular production level. However, the increased IL-21 expression by sJIA T cells was consistent over almost all differentiating conditions (**Figure 2B, C-E**: middle panels; **S3C)** and remained in reciprocal association with IFNγ release, albeit below significance level (**Figure 2 C-E**: right panels). In part consistent with our cellular expression data (**Figure S2 B, C)** we did not detect elevation in IL-17A or IL-22 release in any of the tested conditions, unless present in the respective stimulation cocktail (**Figure S3A, C)**.

**Figure 2.**
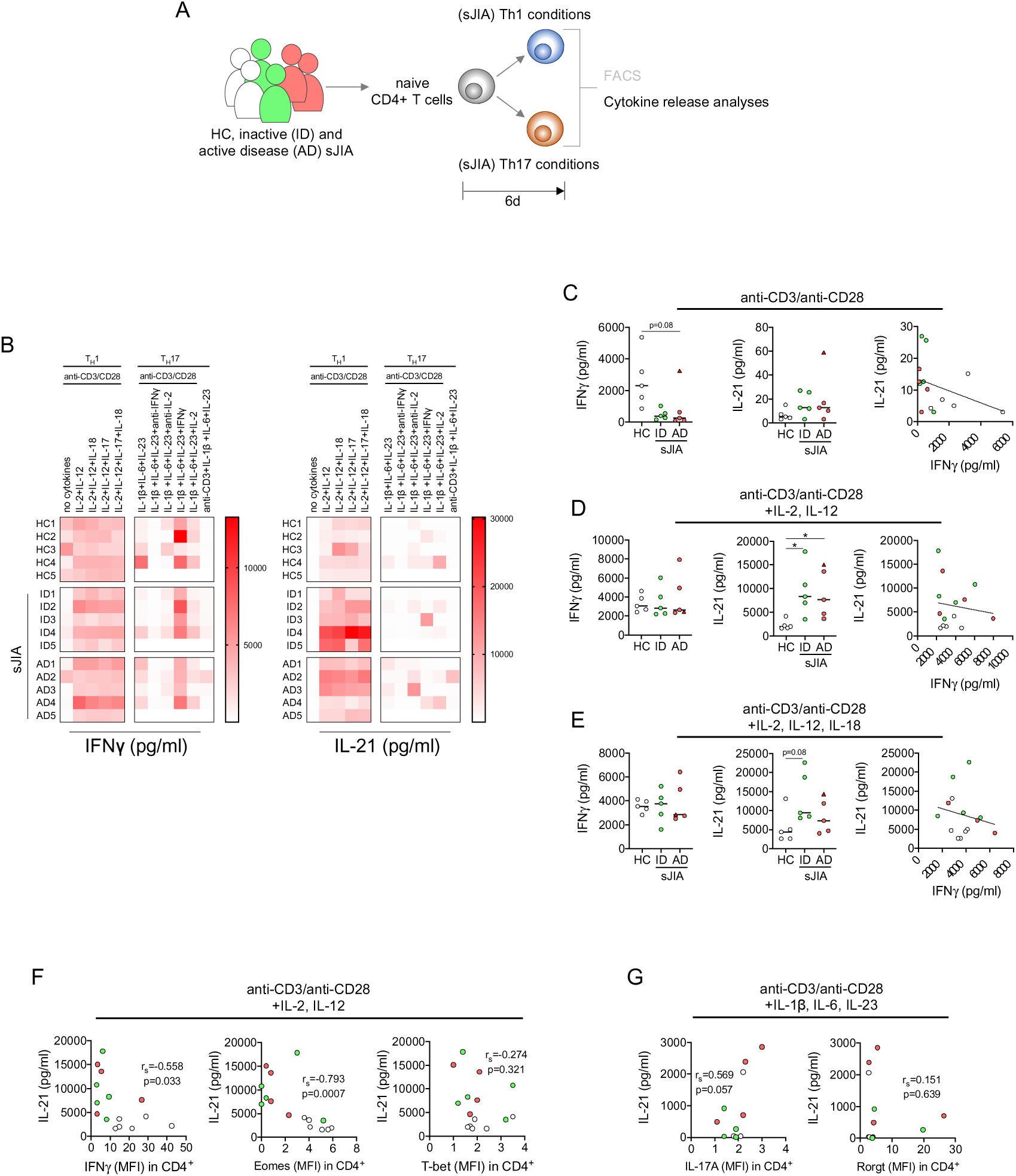
SJIA naïve CD4+ T helper (Th) cell differentiation towards Th1 is hallmarked by IL-21 release in negative association with IFNγ and Eomesodermin expression. **A**, Naive Th cells were isolated from pediatric healthy controls (HC, n=7), active disease (AD, n=5), and inactive disease (ID, n=8) sJIA patients (see table 1) and cultured under T0 (anti-CD3/anti-CD28) and different Th1 and Th17 polarizing conditions. IL-17A and IL-18 were included to mimic a sJIA-cytokine milieu. On day six of culture, cytokine release (IFNγ, IL-17A, IL-21, IL-22) into culture supernatants following phorbol 12-myristate 13-acetate/ionomycin super-stimulation was analyzed by multiplexed bead array assay. **B,** Heatmaps of IFNγ (left panel) and IL-21 (right panel) release (pg/ml) by naive CD4+ cells following culture under the indicated T0, Th1, and Th17 polarizing conditions and super-stimulation as described. **C**-**E**, IFNγ (left panels) and IL-21 (middle panels) release and IFNγ−IL-21 correlation (right panels) of naive CD4+ cells cultured under the indicated Th0 and Th1 polarizing conditions and following super-stimulation. Data is shown in scatter dot plots of the measured cytokine concentrations in pg/ml. The triangular data point refers to a patient who developed MAS subsequent to the analysis. **F**, Correlations of IL-21 release (pg/ml) with intracellular IFNγ (left panel), Eomesodermin (Eomes) (middle panel) and T-bet (right panel) expression (geometric mean fluorescence intensities (MFIs) normalized to unstimulated samples) by cells cultured under the indicated Th1 polarizing condition and super-stimulated. **G**, Correlation of IL-21 release (pg/ml) and intracellular IL-17A (left panel) and RORγt (right panel) expression (MFI) by naïve CD4+ Th cells upon culture under the indicated Th17 polarizing condition and following super-stimulation. **B**-**G**, Each data point presents a value derived from an individual patient or HC. **C**–**E,** Lines indicate median values. Data in left and middle panels were analyzed by Kruskal-Wallis followed by Dunn’s post hoc test; * = *P* < 0.05. **C**-**G**, All correlation data were analyzed for simple linear regression (OLS) and F-test; r_s_ (r-squared value) and *P*-values are indicated.

IL-21 is produced by NKT cells as well as several CD4+ T cellular subsets. Importantly, IL-21 plays a pivotal role in peripheral T cell differentiation(40) and can inhibit IFNγ production in developing Th1 cells through Eomesodermin repression(41). As our flow cytometry data already indicated a tight correlation of IFNγ and Eomesodermin expression in course of Th1 differentiation, we now tested the association of IL-21 release with cellular IFNγ and Eomesodermin expression (**Figure 2F, G**). Among experiments for which both cytokine release and flow cytometry data were available, we observed a marked negative correlation of IL-21 in culture supernatants with both IFNγ and Eomesodermin but not T-bet expression (**Figure 2F**). Further, we noted that recombinant IL-21 present during Th1 differentiation of HD naïve CD4+ T cells to significantly reduce IFNγ expression (**Figure S4**). In addition, we observed a positive association of IL-21 release with cellular IL-17 but not RORγt production among designated Th17 polarizing conditions (**Figure 2G**).

Collectively our data suggest elevated IL-21 expression by sJIA T cells impairs IFNγ-production in developing Th1 cells via Eomesodermin repression.

### Early Th1 differentiation of sJIA naive peripheral CD4+ T cells is shifted toward a Tf/ph phenotype

Among CD4+ T helper cell subtypes T follicular helper (Tfh) cells have been reported to produce IL-21 at particularly high levels. Besides high IL-21 release, Tfh cells are hallmarked by cell surface expression of PD-1, ICOS and CXCR5(42), while a peripheral/tissue-resident counterpart arising in context of chronic inflammation (peripheral T helper (Tph) cells) has been reported to express no or only marginal surface levels of CXCR5(43–45).

Our objective for the next set of experiments was to investigate whether - in the tested conditions - naïve sJIA Th cells might differentiate toward Tf/ph rather than Th1 cells. Therefore, naïve CD4+ T cells from HCs (n=5) and active (n=3) as well as disease-inactive (n=4) sJIA patients (**Table 1**) were driven toward either Th1 or Tf/ph polarization by the indicated conditions (**Figure 3 A, B**). Subsequently, cells were super-stimulated using PMA/ionomycin and were analyzed for expression of IFNγ, IL-21, PD-1, ICOS and CXCR5 by flow cytometry. Among all tested conditions and albeit cytokine expression levels were generally lower in these experiments compared to previous results (**Figure 1**), we again observed the pattern of elevated IFNγ expression by HCs versus sJIA CD4+ T cells (**Figure 3 B, D**, **Figure S5**). In contrast, we noted particularly naïve CD4+ T cells isolated from inactive sJIA patients to overexpress IL-21, PD-1 and ICOS compared to both active disease patients and HCs (**Figure 3B, D**; **Suppl. Figure S5**). IFNγ and IL-21 expression were markedly negatively associated among naïve CD4+ T cells stimulated only by CD3/CD28-ligation (**Figure 3B-D**). This association was weaker or lost in course of Th1 differentiation but partly restored upon Tf/ph polarization (**Figure 3C, D**; **S5**). Further, intracellular IL-21 production was strongly correlated with cell surface expression of PD-1 throughout (**Figure 3C, D**; **S5**). Marked association of IL-21 production and ICOS expression on cells was particularly evident in course of both Th1 and Tf/ph differentiation (**Figure 3C**). In contrast to high PD-1 and ICOS, we observed only marginal CXCR5 expression throughout (**Figure 3B, C**). Besides surface marker and intracellular cytokine expression we also quantified the release of IL-21 and CXCL13 as hallmark molecules produced by both follicular and peripheral T helper cells(45). We observed elevated release of both IL-21 and CXCL13 particularly among cells derived from inactive sJIA patients’ naïve CD4+ T cells (**Figure S6**).

**Figure 3.**
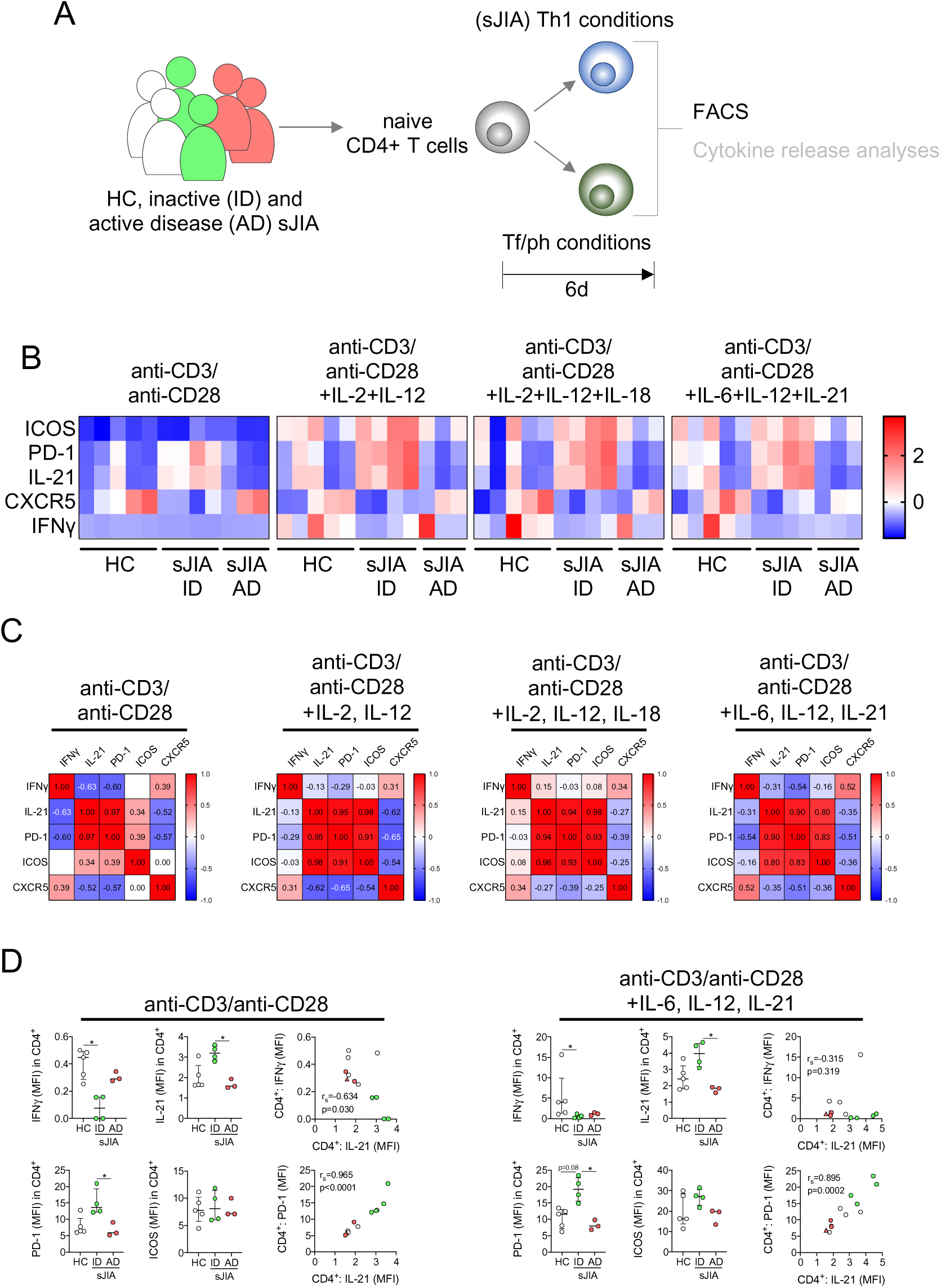
Early CD4+ T helper (Th) cell differentiation in systemic juvenile idiopathic arthritis (sJIA) is shifted toward a (PD-1+ICOS+CXCR5-) peripheral T helper (Tph) cell phenotype. **A**, Naïve Th cells were isolated from pediatric healthy controls (HC, n=5), active disease (AD, n=3), and inactive disease (ID, n=4) sJIA patients (see table 1) were cultured under T0, Th1, and T follicular helper (Tfh) / peripheral T helper (Tf/ph) polarizing conditions. Following six days of culture and super-stimulation with phorbol 12-myristate 13- acetate/ionomycin (4h), cells were analyzed by flow cytometry. **B**, Heatmap of z-scores indicating ICOS, PD-1, IL-21, CXCR5, and IFNγ expression (geometric mean fluorescence intensity (MFI) normalized to unstimulated samples) by naïve CD4+ T cells arising from the indicated Th0, Th1, and Tf/ph polarizing conditions and following super-stimulation. **C**, Multiple correlation analyses of IFNγ, IL-21, PD-1, ICOS, and CXCR5 expression (MFI)as shown in **B**. Spearman’s rank correlation coefficients are indicated. **D**, IFNγ, IL-21, PD-1, and ICOS expression (MFI, left and middle columns) and correlation of IL-21 and IFNγ or PD-1 expression (MFI, right columns) by naïve CD4+ T cells cultured under the indicated Th0 (left panel) and Tf/ph polarizing (right panel) conditions and following super-stimulation. **B**-**D**, Each data point presents a value derived from an individual patient or HC. **D**, Lines indicate median values. Data in left and middle columns were analyzed by Kruskal-Wallis followed by Dunn’s post hoc test; * = *P* < 0.05. Correlation data were analyzed by simple linear regression (OLS) and F-test; r_s_ (r-squared value) and *P*-values are indicated.

Paralleling *in vitro* experiments, we also performed *ex vivo* analyses of patients’ biomaterial. We observed elevated levels of IL-21 as well as IL-6 and IL-23 as key cytokines involved in Tf/ph differentiation in active sJIA patients (**Figure 4A**). Further, in PBMC samples of patients with sJIA we observed decreased Th cellular IFNγ but elevated IL-21 expression, particularly among CD4+PD-1+ and CD4+CXCR5+ cells in inactive sJIA patients’ samples, when compared to pediatric controls (**Figure 4B**). In these samples IL-21 production was best associated with CXCR5 expression (**Figure 4C**). Importantly, we also analyzed already available whole blood RNA sequencing data (GSE112057(46)) obtained from healthy controls (n=12) and sJIA (n=26) as well as polyarticular (n=46) and oligoarticular JIA patients (n=43) for sJIA-hallmark (*IL1B*, *IL1R1*, *IL1RN*, *S100A12*), Tph (*CX3CR1*, *CXCR6*, *CCR2*, *CCR5*, *PRDM1*), Tfh (*FOS*, *CXCR5*, *BCL6*), naïve (*LRRN2*, *CCR7*, *NPM1*), central memory (*PASK*), effector memory (*IL7R*, *KLRB1*, *TNFSF13B*) and activated effector memory (*CCL4*, *GZMH*, *GZMA*) T cell signature gene expression(47, 48). Beyond, we also looked at *IFNG* as well as *TBX21* (T-bet), *EOMES*, *MAF*, and *SOX4* transcription (**Figure 4D**). Expression of most analyzed genes, including *IFNG*, was decreased in all JIA entities when compared to healthy controls (**Figure 4D, F**). However, alongside with elevated expression of *IL1B*, *IL1R1*, *IL1RN* and *S100A12* as hallmarks for sJIA (**Figure 4D, E**), we also observed strongly elevated expression of *BCL6* as well as *TNFSF13B*, and *FOS* expression elevated by trend (**Figure 4D, F**).

**Figure 4.**
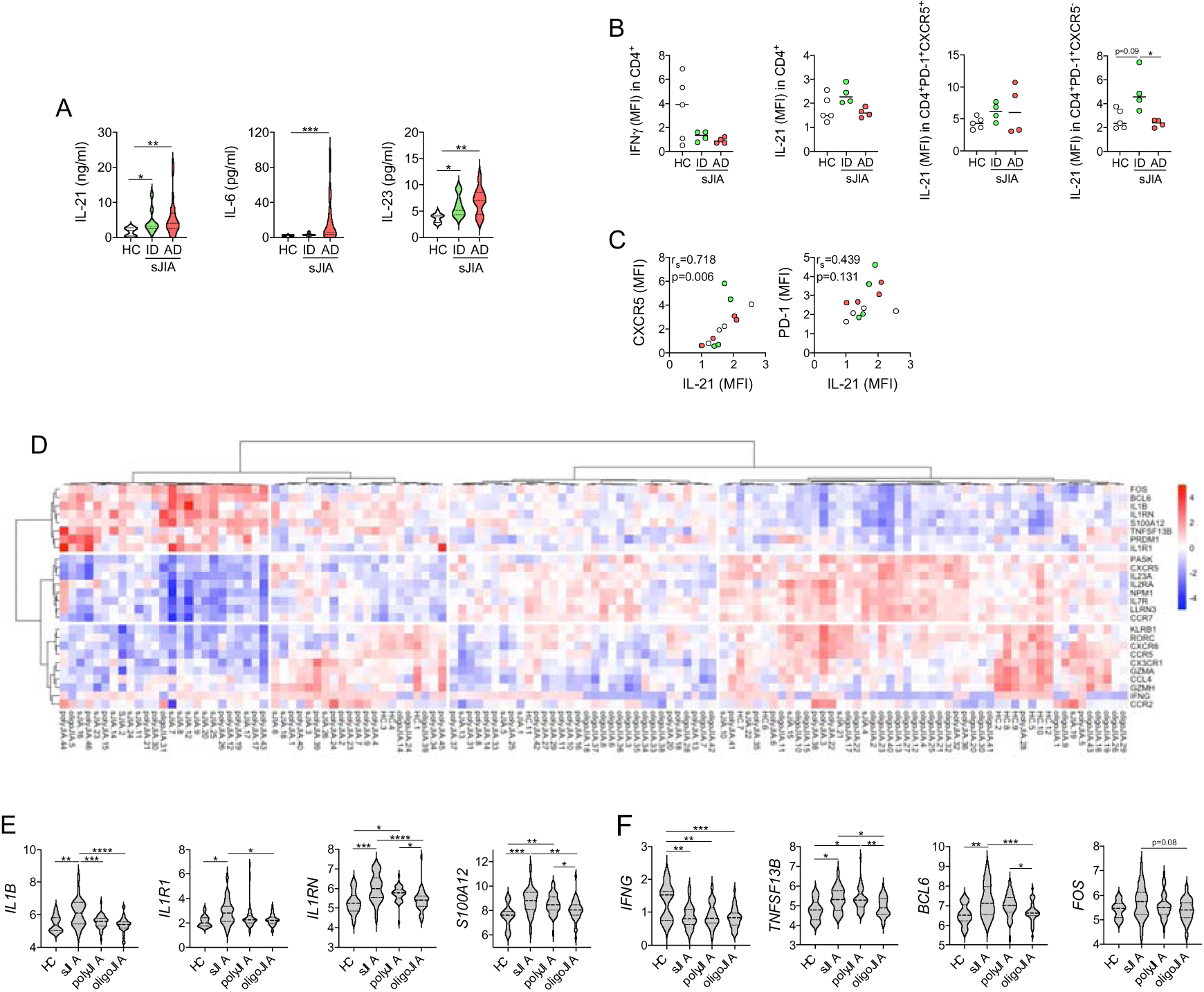
Circulating CD4+ T helper (cTh) cells in systemic juvenile idiopathic arthritis (sJIA) reveal a T follicular helper (Tfh) / peripheral T helper (Tph) cell signature. **A**, IL-21 (left), IL-6 (middle), and IL-23 (right) serum levels in pediatric healthy controls (HC, n=10), active disease (AD, n=27), and inactive disease (ID, n=9) sJIA patients (see table 1). Data is shown in violin plots of the measured cytokine concentrations in pg/ml. **B,** Flow cytometry analyses of IFNγ and IL-21 in indicated cTh cell subsets in peripheral blood mononuclear cells (PBMCs) isolated from pediatric HCs (n=5), AD (n=3), and ID (n=4) sJIA patients (see table 1) following super-stimulation with phorbol 12-myristate 13-acetate/ionomycin (4h). **C**, Correlation of IL-21 with CXCR5 (left panel) and PD-1 expression (geometric mean fluorescence intensities (MFIs) normalized to unstimulated samples, right panels) in cTh cells. **D**, Heatmap of unsupervised clustering using correlation distance and ward.D linkage of indicated gene expression in whole blood RNA sequencing data (GSE112057(46)) from HC (n=12) as well as sJIA (n=26), polyarticular (n=46) (polyJIA) and oligoarticular JIA (oligoJIA) patients (n=43). **E** and **F**, Expression of SJIA-hallmark genes (**E**, *IL1B*, *IL1R1*, *IL1RN*, *S100A12*) as well as (**F**) reported T cell associated genes *IFNG*, *TNFSF13B*) (T effector memory), *BCL6* and *FOS* (Tfh) as in **D** depicted in violin plots as the relative difference in expression of the respective genes. **B**-**D**, Each data point presents a value derived from an individual patient or HC. **A**, **B**, **E**, and **F**, Lines indicate median values. Data were analyzed by Kruskal-Wallis followed by Dunn’s post hoc test; * = *P*<0.05, ** = *P*<0.01, *** = *P*<0.001, **** = *P*<0.0001. **C**, Simple linear regression (OLS) and F-test; r_s_ (r-squared value) and *P*-values are indicated.

Collectively, these data imply Th differentiation in sJIA both *in vitro* and *in vivo* to skew toward a Tf/ph phenotype.

### Tf/ph cells derived from sJIA naïve CD4+ cells are functional and drive B cellular plasma-blast generation

Tfh cells reside within B cell follicles to provide B cell help to produce high-affinity antibodies, while Tph cells are thought to drive corresponding plasma-blast generation in the periphery, namely within inflamed tissue(45). Therefore, we aimed to test whether naïve sJIA CD4+ T cells subjected to Th1 or Tf/ph differentiating conditions would indeed result in functional Tf/ph cells with the ability to drive B cellular plasma-blast generation.

We isolated naïve CD4+ T cells from HCs (n=4) and active (n=2) as well as disease-inactive (n=3) sJIA patients (**Table 1**). Cells were either cultured with anti-CD3/CD28 or driven toward Th1 (anti-CD3/CD28+IL-2+IL-12) or Tf/ph (anti-CD3/CD28+IL-6+IL-12+IL-21) polarization (**Figure 5A**). Subsequently, cells were co-cultured with freshly isolated healthy donor memory B cells for six further days and eventual plasma-blast generation (CD19^+^IgD^-^ CD27^+^CD38^+^CD138^-^) was analyzed by flow cytometry (**Figure 5A**). Plasma blasts were defined as CD19^+^IgD^-^CD27^+^CD38^+^CD138^-^ cells(49) (**Figure 5B-D**). We found that Tf/ph cells generated from naïve sJIA CD4+ B cells using the indicated conditions resulted in a marked increase in the frequency of plasma-blasts compared to cultures with CD3/CD28 ligation without polarizing cytokines (**Figure 5D, E**). This increase was most pronounced with designated Tf/ph differentiating conditions and significantly elevated over healthy controls (**Figure 5D-G**).

**Figure 5.**
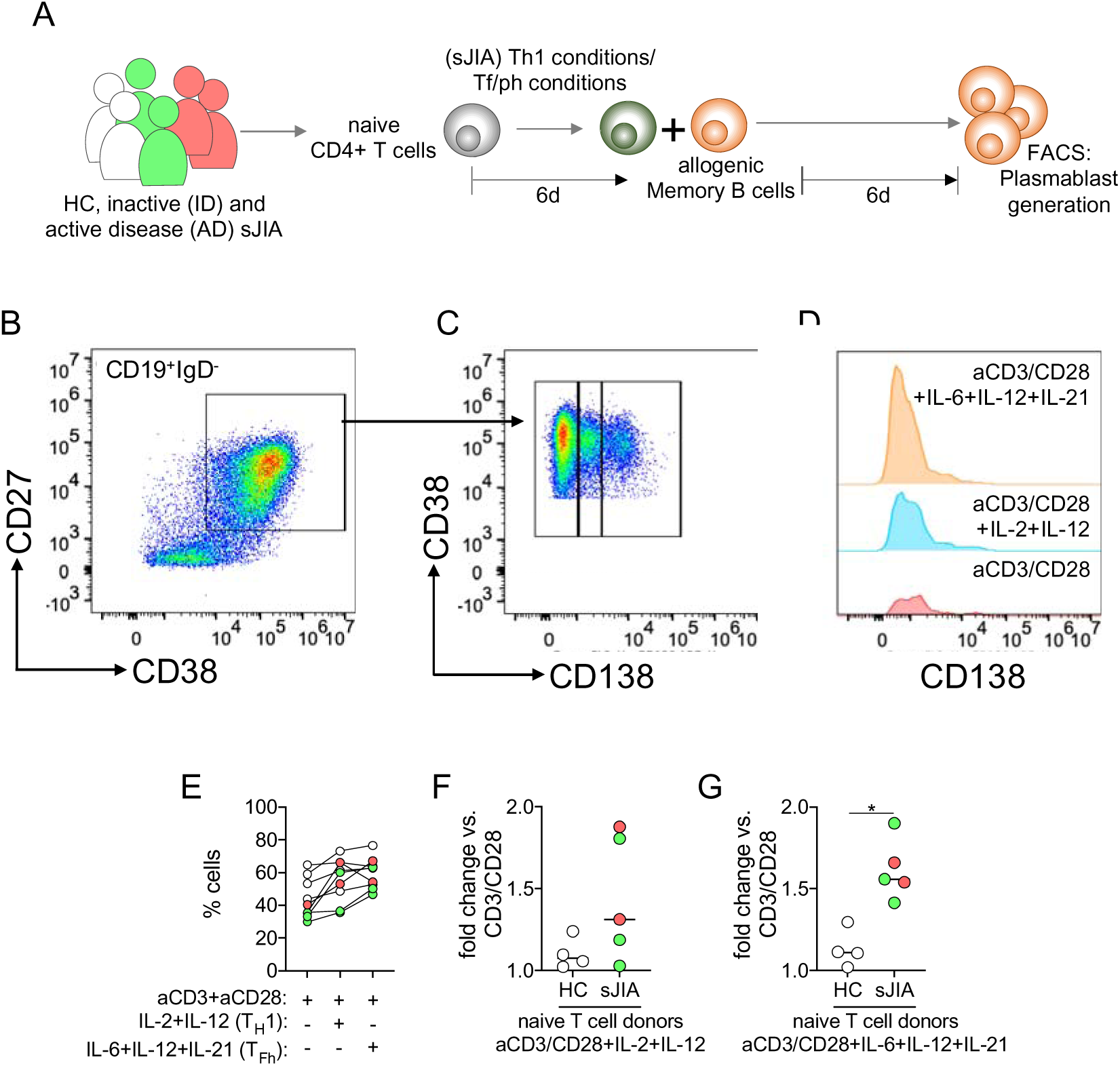
**T follicular helper / peripheral T helper cells (Tf/ph) derived from sJIA naïve CD4+ T helper (Th) cells can drive enhanced B cellular plasma blast generation**. **A,** Naïve Th cells were isolated from pediatric healthy controls (HC, n=4), active disease (AD, n=2), and inactive disease (ID, n=3) sJIA patients (see table 1) and cultured under T0, Th1, and Tf/ph polarizing conditions. Following six days of culture polarized Th cells were co-cultured with allogenic HC memory B cells isolated from a single healthy donor and the generation of plasma blasts was assessed by flow cytometry following another six days of culture. **B-D**, Representative flow cytometry pseudo-color (**B, C**) or histogram plots (D) to monitor plasma blast (CD19+IgD-CD27+CD38+CD138-) generation from co-cultured memory B cells. **E**, Frequency of plasma blasts in co-cultures with Th cells polarized under the indicated Th0, Th1, and Tf/ph polarizing conditions. **F** and **G**, Relative change in the frequency of plasma blasts in co-cultures with Th cells cultured under the indicated Th1 (F) and Tf/ph (G) polarizing conditions, normalized to the Th0. **E-G**, Each data point presents a value derived from an individual patient or HC. F and G, Lines indicate median values. Data were analyzed by Mann-Whitney U test; * = *P*<0.05.

Finally, we also questioned whether an environment of low IFNγ might benefit IL-17A expression from γδT cells, as we previously speculated(18). In these experiments, inflammatory stimulation of γδT cells in co-culture with human monocytes with or without an IFNγ-neutralizing antibody (**Figure S7A**) partly depleted IFNγ (**Figure S7B**) but significantly enhanced the γδT cellular IL-17A release (**Figure S7C**).

Collectively our data suggest that the differentiation of naïve CD4+ T cells in sJIA is committed to differentiation toward functional Tf/ph cells and that an IFNγ-depleted environment may benefit γδT cellular IL-17A release as an independent pathophysiological sJIA feature.

## Discussion

The contribution of adaptive immunity to sJIA pathogenesis is still unclear. Although the disease is classified among polygenic autoinflammatory syndromes, observations such as an association with the MHCII haplotype *HLA-DRB1*11*(15) and Treg polarization towards Th17 in chronic sJIA(16) complicate a clear-cut classification. In this context we previously reported γδT cells in sJIA to operate as main source for IL-17A, while we found peripheral Th cells to express comparably little IL-17A and IFNγ, in particular(18). These observations echoed data from a murine sJIA model, which reported elevated γδT cellular IL-17 expression particularly on IFNγ-/- background(34) as well as reports suggesting limited in vivo cell exposure to circulating IFNγ(33).

In the present study, we report that in sJIA, the differentiation of naïve peripheral Th cells towards a Th1 and IFNγ production is impaired and instead skewed towards a Tf/ph phenotype. This we observed particularly among cells obtained from inactive patients while *in vitro* differentiated cells were hallmarked by elevated IL-21 and CXCL13 production and cell-surface expression of PD-1 and ICOS. *Ex vivo* analyses mirrored *in vitro* data in that we found elevated IL-21 expression both at cellular as well as serum level and identified *BCL6* as a master transcription factor for Tf/ph differentiation(50–52) as highly overexpressed in retrospective analyses of sJIA whole blood RNA sequencing data(46). Finally, *in vitro* generated sJIA Tf/ph cells proofed functional as they could drive plasma blast generation from allogenic memory B cells.

In line with the window-of-opportunity model of sJIA pathogenesis and our as well as others’ previous data, the central aim of our study was to investigate the power of the sJIA cytokine environment on influencing Th cell differentiation. Surprisingly, we observed Th cell differentiation in sJIA to be altered already at naïve T cell level. Even when removed from the individual sJIA patient environment and exposed to controlled *in vitro* stimulating conditions, naïve sJIA T cells appeared wired for Tf/ph differentiation. Both in *in vitro* polarization experiments, as well as in *ex vivo* analyses this was most consistently hallmarked by overexpression of IL-21. Besides Th17 cells, Tf/ph cells are the most prominent producers of IL-21(40). Yet, in our experiments, Th1 differentiating conditions applied on sJIA naïve T cells resulted in massive IL-21 release, while Th17 polarizing stimuli also promoted IL-21 production in positive correlation with IL-17A; yet, this was far less than seen with sJIA T cells driven towards Th1.

Besides high expression of IL-21 and the B cell chemoattractant CXCL13(53), Tfh cells are hallmarked by PD-1, ICOS and CXCR5 expression. However, in developing sJIA Th cells cultured under Th1 or Tf/ph polarizing conditions *in vitro*, we only detected marginal CXCR5 expression. The observed PD-1+ICOS+CXCR5- phenotype is rather characteristic for Tph cells, a subset of Th cells that has recently been described to activate memory B cells within inflamed tissues and contribute to the formation of tertiary lymphoid organs (TLO) in different autoimmune conditions(43–45).

*Ex vivo*, we found both CD4+PD-1+CXCR5+ and CD4+PD-1+CXCR5-cells to reveal increased cellular IL-21 production, which particularly correlated with CXCR5 expression. In addition, retrospective analyses of whole blood RNA sequencing data (GSE112057(46)) demonstrated an sJIA-specific overexpression of the Tfh master transcription factor Bcl-6(50–52). Importantly, whole transcriptome transcription factor motif enrichment analyses previously reported Bcl-6 to be amongst the most prominent binding motifs in sJIA immune cells(54). In contrast, we found no relevant elevation of transcription factor expression such as of Sox-4 (*SOX4*) or Blimp-1 (*PRDM1*)(43–45) or of other genes (*CX3CR1*, *CXCR6*, *CCR2*, *CCR5*)(47) reported to rather associate with Tph differentiation.

Comparing our *in vitro* and *in vivo* data, it might appear that *in vivo*, sJIA Th cells are preferentially skewed toward a Tfh and to a lesser extent towards a Tph cell phenotype as observed in *in vitro* experiments. However, most likely, Tfh and Tph cells co-evolve from a common naïve Tf/ph precursor cell in secondary lymphoid organs (SLOs) under the influence of stimuli favoring the polarization towards both lineages. The tipping point that eventually defines a Tfh or Tph fate is yet unknown(45). Further, Tph cell detection in the circulation may be challenging, as particularly in inflammatory conditions Tph cells mature and reside in inflamed tissue. In addition, immature circulating Tph (cTph) cells presumably express CXCR5 at low levels, which makes them hardly distinguishable from circulating Tfh (cTfh) cells(45). Collectively, this can explain why our *in vivo* data point towards a Tfh dominance in circulation, while our *in vitro* experiments mimicking the pro-inflammatory cytokine environment at the site of inflammation may rather give rise to cells of a Tph phenotype. Regardless of these subtle differences, our data suggest that in sJIA naïve Th cell differentiation is wired for a Tf/ph precursor phenotype that in dependency on localization and stimuli can mature into either Tfh cells in SLOs or into Tph cells at sites of inflammation.

The precise molecular mechanisms that result in a skewing of naïve sJIA Th cell differentiation toward a Tf/ph phenotype remain yet elusive. Tf/ph polarizing stimuli include activin A, IL-12, IL-23, TGFβ and IL-6, that via STAT3 and STAT4 signaling promote the expression of IL-21 and IL-21R, which in turn fosters Tfh differentiation in an autocrine manner(55, 56). We and others observed elevated serum levels of IL-23 and IL-6 in sJIA that might favor Tf/ph polarization and increased STAT3 and STAT4 signaling as well as elevated IL-23R expression in sJIA patients has been reported elsewhere(54). Hence, Tf/ph polarization in sJIA might be incited by elevated levels of Tf/ph polarizing cytokines. Another crucial factor that favors Tf/ph differentiation over Th1 differentiation is the oppression of Eomesodermin expression(57). Importantly, IL-21 has been described to impair Eomesodermin expression and thereby limit IFNγ production in developing Th1 cells(41). In our study, negative correlation of IL-21 release with cellular IFNγ and Eomesodermin expression as well as proof-of-principle data of recombinant IL-21 impairing IFNγ production from Th1 polarized HC naïve T cells support this concept. Hence, developing sJIA Tfh/Tph cells might stabilize their phenotype in an autocrine manner via the release of IL-21, that in turn impairs Eomesodermin and IFNγ expression.

Of note, in the *in vitro* differentiation experiments we observed the Tph phenotype in developing Th cells to arise particularly from naïve T cells isolated from inactive sJIA patients. This was mirrored by *ex vivo* observations. While in the *ex vivo* setting this may reflect that cells in active patients preferentially move to inflamed tissue rather than to reside in the periphery, the enhanced wiring toward *in vitro* Tph differentiation of naïve T cells obtained from inactive *versus* active sJJA patients is puzzling. Interestingly, we observed that the addition of IL-18 as a cytokine highly overexpressed in active sJIA(58) as well as IL-17A partly restored the expression of IFNγ and Eomes in developing active but not inactive sJIA patients’ Th cells. Hence, we speculate that naïve Th cells isolated from active patients originate from an environment of high IL-18 as well as other pro-inflammatory cytokines, and consequently might be less prone to impaired Th1 cell differentiation. Yet, further studies are required to define whether these data indicate a rescued Th1 phenotype or the polarization toward a subset of IFNγ-expressing Tph/Tfh cells, as reported in context of rheumatoid arthritis(59). Nonetheless, regardless of disease activity status we observed elevated IL-21 release and a decreased IFNγ and Eomesodermin expression in developing Th cells even if cells underwent no stimulation other than by CD3/CD28-ligation. In summary, this indicates a preferential differentiation toward a Tf/ph over a Th1 phenotype as a general pattern in sJIA. Intriguingly, in oligo- and polyarticular JIA naïve T cell differentiation toward Th1 was reported to rather skew toward an inflammatory effector Th1.17 and Th17 subtype(60). In light of our present data, this suggests aberrant Th cell polarization as a general phenomenon in both systemic and non-systemic JIA.

Nonetheless, our observations must be interpreted in the light of three main limitations. 1) Most of our experiments comprise relatively limited patient numbers and we did not have access to cells from other compartments than the periphery. Yet, we used independent patient cohorts for the initial Th1/Th17 differentiation and subsequent Th1/Tfh polarization experiments, which were consistent in main study findings such as elevated IL-21 but decreased IFNγ expression. Further, retrospective analyses of whole blood RNA sequencing data revealing sJIA-specific *BCL6* overexpression strengthen our data on a different level. 2) We did not harmonize patients for specific disease activity features as well as disease duration and we cannot rule out an underlying confounding effect on our observations. 3) Finally, even though we demonstrate functionality of the Tf/ph cells arising from naïve sJIA T cells in terms of driving B cellular plasma blast generation, we cannot yet pinpoint their specific contribution to sJIA pathogenesis or progression. Existing data on presence of antinuclear antibodies and rheumatoid factor in arthritic sJIA patients are still limited to small overall patient numbers(61).

Despite these limitations, we believe our observations to contribute to the understanding of sJIA pathogenesis, suggesting a role of adaptive immune cells, namely Tf/ph cells in disease, in line with the bi-phasic model of disease progression(3, 6). Intriguingly, as an important cytokine in Tf/ph polarization, IL-6 targeting therapies, which are considered second-line therapeutic options in refractory arthritic disease, might interfere with respective Th cell differentiation. Longitudinal data collection could define the impact of cytokine blocking therapies on the Tf/ph compartment in sJIA. Of note, treatments specifically blocking Tf/ph polarization by interrupting IL-21 signaling have proved safe and effective in first clinical trials in rheumatoid arthritis(62).

## Data Availability

All data produced in the present study are available from the corresponding author upon reasonable request.

## Author contributions

JK, SS, CP and CK acquired laboratory data; AH, CH, HW, DH and DF collected and analyzed clinical data; JK, DF and CK designed the study; Data analysis & interpretation: all authors; JK and CK wrote the manuscript; Manuscript draft & revision: all authors

## Supplementary methods

### Human peripheral blood mononuclear cell (PBMC) isolation

PBMCs were isolated by density-gradient centrifugation (Pancoll, density: 1.077 g/ml, PAN-Biotech GmbH, Germany) from peripheral blood samples collected from sJIA patients and pediatric HCs or residual blood obtained from adult healthy thrombocyte donors. Until further assessment, the cells were frozen at -150 °C in 10 % dimethyl sulfoxide (DMSO; Carl Roth GmbH + Co. KG, Germany) / 90 % fetal calf serum (FCS; Gibco^TM^, Thermo Fisher Scientific Inc., Waltham, MA, USA) at 1 x 10^7^ cells x ml^-1^. For analysis, the PBMCs were thawed in RPMI 1640 medium supplemented with 10 U Penicillin / 0.1 mg x ml^-1^ Streptomycin, 2 mM L-glutamine, 1 x non-essential amino acids, 1 mM sodium pyruvate (all purchased from Merck KGaA, Germany), and 10 % FCS. The PBMCs obtained from one study sample were used for both an *ad-hoc* flow cytometry analysis of Th cell markers at aliquots of 1 x 10^5^ cells per sample, and for the isolation of naïve Th cells for an *in vitro* characterization of Th cell differentiation.

### Isolation, culture, and stimulation of naïve T helper (Th) cells

Naïve Th cells were isolated from PBMCs by immunomagnetic negative selection using the EasySep^TM^ Human Naïve CD4+ T Cell Isolation Kit II (STEMCELL Technologies Inc., Cologne, Germany) according to the manufacturer’s instruction. Naïve Th cells were diluted to 5 x 10^5^ cells x ml^-1^ in supplemented RPMI 1640 medium and plated at a density of 5 x 10^4^ cells / well in 96-well U-bottom suspension culture plates (Greiner Bio-One, Frickenhausen, Germany) coated with α-human CD3 (5 µg x ml, clone: OKT3, BioLegend, San Diego, CA, USA). Cells were differentiated for 6 days (37°C, 5% CO_2_) in the presence of combinations of the following cytokines, stimuli, or blocking agents, as indicated and detailed in the respective results: α-human CD28 (2.5 µg x ml^-1^, clone: CD28.2, BioLegend), α-human IFNγ (1 µg x ml^-1^, clone: NIB42, Thermo Fisher Scientific Inc.), α-human IL-2 (10 µg x ml^-1^, clone: MQ1-17H12, BioLegend,), recombinant human IFNγ (1000 U x ml^-1^, PeproTech, Hamburg, Germany), recombinant human IL-1β (50 ng x ml^-1^, BioLegend), recombinant human IL-2 (100 U x ml^-1^, PeproTech), recombinant human IL-6 (5 ng x ml^-1^, PeproTech), recombinant human IL-12 (5 ng x ml^-1^, PeproTech), recombinant human IL-17A (10 ng x ml^- 1^, BioLegend), recombinant human IL-18 (5 ng x ml^-1^, Invivogen, San Diego, CA, USA), recombinant human IL-21 (10 ng x ml^-1^, BioLegend), and recombinant human IL-23 (50 ng x ml^-1^, BioLegend).

### Co-cultures of Th cells and allogenic memory B cell

Co-cultures were set up as described recently(43). Naïve Th cells were isolated from PBMCs and cultured for six days at conditions indicated in the present study and according to methods and concentrations as described above. Following six days of culture cells were harvested, washed twice with cell culture medium (RPMI), and were counted. Memory B cells were isolated from PBMCs obtained from a single healthy donor using the EasySepTM Human Memory B Cell Isolation Kit (STEMCELL Technologies Inc.) according to the manufacturer’s instructions. Polarized T cells and isolated memory B cells were co-cultured at a 1:10 ratio (range of T cells numbers used in cultures: 2800-13000) in the presence of recombinant *Staphylococcus aureus* enterotoxin B (1 µg x ml^-1^, Hoelzel Diagnostika, Cologne, Germany) and LPS (5 µg x ml^-1^, Lipopolysaccharides from *Escherichia coli* O55:B5, Sigma-Aldrich, Taufkirchen, Germany). Following six days of co-culture (37°C, 5% CO_2_), the cells were harvested, washed, and prepared for flow cytometry staining.

### Flow cytometry

Before flow cytometry analysis, PBMCs and Th cells were stimulated by incubation with phorbol 12-myristate 13-acetate (PMA) and ionomycin (eBioscience^TM^ Invitrogen^TM^ Cell Stimulation Cocktail, Thermo Fisher Scientific Inc.) in the presence of Brefeldin A and Monensin (eBioscience^TM^ Invitrogen^TM^ Protein Transport Inhibitor Cocktail, Thermo Fisher Scientific Inc.) for 5 hours at 37 °C, or left unstimulated. For cell surface marker staining, 1×10^5^ cells were incubated with the respective antibody at a concentration of 1 µl / 100 µl sample in a buffer containing 1 x phosphate-buffered saline (PBS; Merck KGaA, Darmstadt, Germany), 0.1 % bovine serum albumin (BSA; Carl Roth GmbH, Karlsruhe, Germany), 2 mM ethylenediaminetetraacetic acid (EDTA; Carl Roth GmbH), and 0.05 % sodium azide (Merck KGaA) for 30 minutes at 4 °C in the dark.

The following antibodies were used for T cell surface marker staining: α-human CD3-APC-Cy7 (clone: OKT3, BioLegend), α-human CD4-APC (clone: OKT4, BioLegend), α-human CD4-BV510 (clone: OKT4, BioLegend), α-human CD161-PE-Dazzle (clone: HP-3G10, BioLegend), α-human CD183/CXCR3-APC-Cy7 (clone: G025H7, BioLegend), α uman CD185/CXCR5-PE-Dazzle (clone: J252D4, BioLegend), α-human CD196/CCR6-BV650 (clone: G034E3, BioLegend), α-human CD278/ICOS-FITC (clone: C398.4A, BioLegend), and α human CD279/PD-1-BV650 (clone: EH12.2H7, BioLegend).

The following antibodies were used for B cellular plasmablast surface marker staining: CD19-PE-CF594 (clone: HIB19, BioLegend), CD20-FITC (clone: 2H7, BioLegend), CD27-PE-Fire780 (PC7, clone: O323, BioLegend), CD38-BV421 (PB450, clone: HB-7, BioLegend), CD138-APC (clone: DL-101, BioLegend), and IgD-AF7000 (APC-A700, clone: IA6-2, BioLegend).

For the subsequent staining of intracellular cytokines and transcription factors, the cells were fixed and permeabilized by incubation in FixPerm Buffer (eBioscience^TM^ Fixation/Permeabilization Concentrate, eBiosciene^TM^ Fixation/Permeabilization Diluent, Thermo Fisher Scientific Inc., USA) for 30 minutes at room temperature. For intracellular staining, 1×10^5^ cells were incubated with the respective antibody at a concentration of 1 µl / 100 µl sample in 1 x Permeabilization Buffer (eBioscience^TM^, Thermo Fisher Scientific Inc., USA) for 30 minutes at 4 °C in the dark. The following antibodies were used for intracellular staining: α-human Eomesodermin (EOMES)-PE-Cy7 (clone: WD1928, Thermo Fisher Scientific Inc., USA), α-human IFNγ-BV421 (clone: 4S.B3, BioLegend, USA), α-human IL-17A-PE (clone: BL168, BioLegend, USA), α-human IL-21-PE (clone: 3A3-N2, BioLegend, USA), α-human RORγt-APC (clone: AFKJS-9, Thermo Fisher Scientific Inc., USA), and α- human T-bet-AF488 (clone: 4B10, BioLegend, USA). Fluorescence intensities were measured with CytoFLEX S (Beckman Coulter, USA). For compensation and gating, fluorescent beads and unstained samples were used, respectively. Measurement settings were not changed throughout the study. Cytometric data were analyzed by FlowJo (version: v10.0.8, Becton, Dickinson and Company, USA). Expression levels of cytokines and transcription factors were assessed as geometric mean fluorescence intensities (MFIs). MFIs were normalized to unstained samples, and in a second step, to control differentiation conditions (α-CD3 or α-CD3 and α-CD28 only).

### Multiplexed bead array assays

Reagents for multiplexed quantification of CXCL13, IFNγ, IL-17A, IL-21, and IL-22 in supernatants of the Th cell cultures as well as of IL-6, IL-21, and IL-23 in sera were purchased from R&D Systems (Minneapolis, OH, USA). Reagents and sera or cell culture supernatants were prepared according to the manufacturer’s instructions (R&D Systems). Data acquisition and analysis were performed on a MAGPIX instrument (Merck Millipore, Darmstadt, Germany) using xPONENT v4.2 software (Luminex).

### γδT cell-monocyte co-cultures

Co-cultures of γδT cells and monocytes were set up as described previously(18). Briefly, HC γδ T cells were isolated from PBMCs prepared from residual blood obtained from thrombocyte donors by immunomagnetic negative selection using the EasySep™ Human γ/δ T Cell Isolation Kit (STEMCELL Technologies Inc.) according to the manufacturer’s instructions. Monocytes were prepared from fresh HC whole blood using the EasySep™ Human Monocyte Isolation Kit (STEMCELL Technologies Inc.) according to the manufacturer’s instructions. γδT cells and monocytes were co-cultured at a ratio of 1:2 in RPMI 1640 medium supplemented with the phosphoantigen isopentenyl pyrophosphate (IPP; 10 μg x ml ; Merck KGaA), or the nitrogen-containing bisphosphonates zoledronate (1μM x ml^-1^, Sigma-Aldrich) for six days. In case of stimulations with zoledronate, monocytes were cultured with the indicated concentration for 2 hours, then were washed twice with RPMI, and subsequently co-cultured with γδT cells. All co-cultures were supplemented with recombinant human IL-2 (100 U x ml^-1^, PeproTech) and further stimulated with indicated combinations of recombinant human IL-1β (50 ng x ml, BioLegend), recombinant human IL-18 (5 ng x ml, Invivogen), and recombinant human IL-23 (50 ng x ml^-1^, BioLegend), as well as anti-human IFN γ (1 µg x ml^-1^, clone: NIB42, Thermo Fisher Scientific Inc.),

### ELISA

IFNγ release by healthy donor Th cells in experiments +/- recombinant IL-21, as well as IFNγ concentration and IL-17A release in γδT cell-monocyte co-cultures, were determined by ELISA. Cell culture supernatants following six days of culture in the indicated conditions and super-stimulation with phorbol 12-myristate 13-acetate/ionomycin were collected after centrifugation and frozen at -20°C until analysis. For quantification of the IFNγ and IL-17A concentrations in the supernatants, ELISA MAX^TM^ kits (BioLegend) were used according to the manufacturer’s instructions. Quantification was performed in Nunc™ 96-well polypropylene microplates (Thermo Fisher Scientific Inc.). The plates were washed with a buffer containing 140 mM NaCl, 4 mM KH_2_PO_4_, 12 mM Na_2_HPO_4_ x 2 H_2_O, and 0.04 % (v/v) Tween® 20. Per sample, the assay was performed in 2 technical replicates, of which the mean IFNγ and IL-17A concentrations were calculated.

### Statistical analyses

For statistical analyses, the statistics software GraphPad Prism for Mac OS X was used (version 6.0f, GraphPad Software Inc., USA). To test for a linear correlation between the expression of cell surface markers, transcription factors, and cytokines in restimulated PBMCs and Th cells, a simple linear regression model was fit to the data using the OLS method. If a significant association was found using an F test, the r-squared value (r_s_) and the p-value are indicated. To analyze the ELISA and cytokine beads assay results, a non-linear log-log fit of baseline-corrected standard values was applied, and assayed sample values were interpolated from the respective standard curve. To compare median cytokine concentration in patient serum samples and the supernatant of distinct Th cell and γδ T cell differentiation conditions, Kruskal-Wallis-test and post hoc Dunn’s test for nonparametric data were used since all the samples did not pass D’Agostino and Pearson normality test and/or respective skewness was > |1|. Similarly, to compare the median or geometric mean expression of cell surface markers, transcription factors, and cytokines in restimulated PBMCs, Th cells, and γδ T cells, Kruskal-Wallis-test and post hoc Dunn’s test were employed. To compare the median cytokine concentration in the supernatant of two distinct differentiation conditions, the Mann-Whitney U test was employed. Where statistically significant differences were observed, p-values are indicated as follows: * P < 0.05; ** P < 0.01; *** P < 0.001, **** P < 0.0001. Corrected P < 0.05 was considered statistically significant.

---

**Figure S1.**
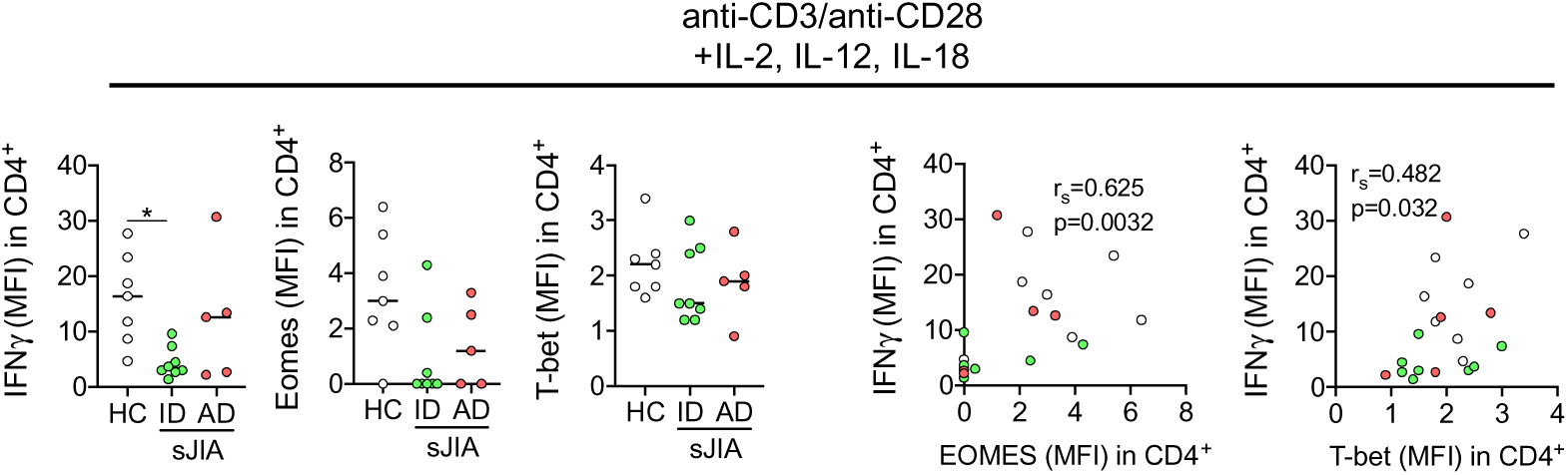
SJIA naïve CD4+ T helper (Th) cell differentiation towards Th1 results in low IFNγ and Eomesodermin expression. Naive Th cells were isolated from pediatric healthy controls (HC, n=7), active disease (AD, n=5), and inactive disease (ID, n=8) sJIA patients (see table 1) and cultured under T0 (anti-CD3/anti-CD28) and different Th1 and Th17 polarizing conditions. Following six days of culture and super-stimulation with phorbol 12- myristate 13-acetate/ionomycin (4h), cells were analyzed by flow cytometry (see **Figure 1**). IFNγ, Eomes, and T-bet expression (geometric mean fluorescence intensity (MFI) normalized to unstimulated samples), as well as correlation of IFNγ with Eomes or T-bet expression (MFI) in cells cultured under indicated conditions. Each data point represents a value derived from an individual patient or HC. Lines in scatter plots indicate median values, data were analyzed by Kruskal-Wallis test followed by Dunn’s post hoc test; * = *P* < 0.05. Correlation data were analyzed by simple linear regression (OLS) and F-test; r_s_ (r-squared value) and *P*-values are indicated.

**Figure S2.**
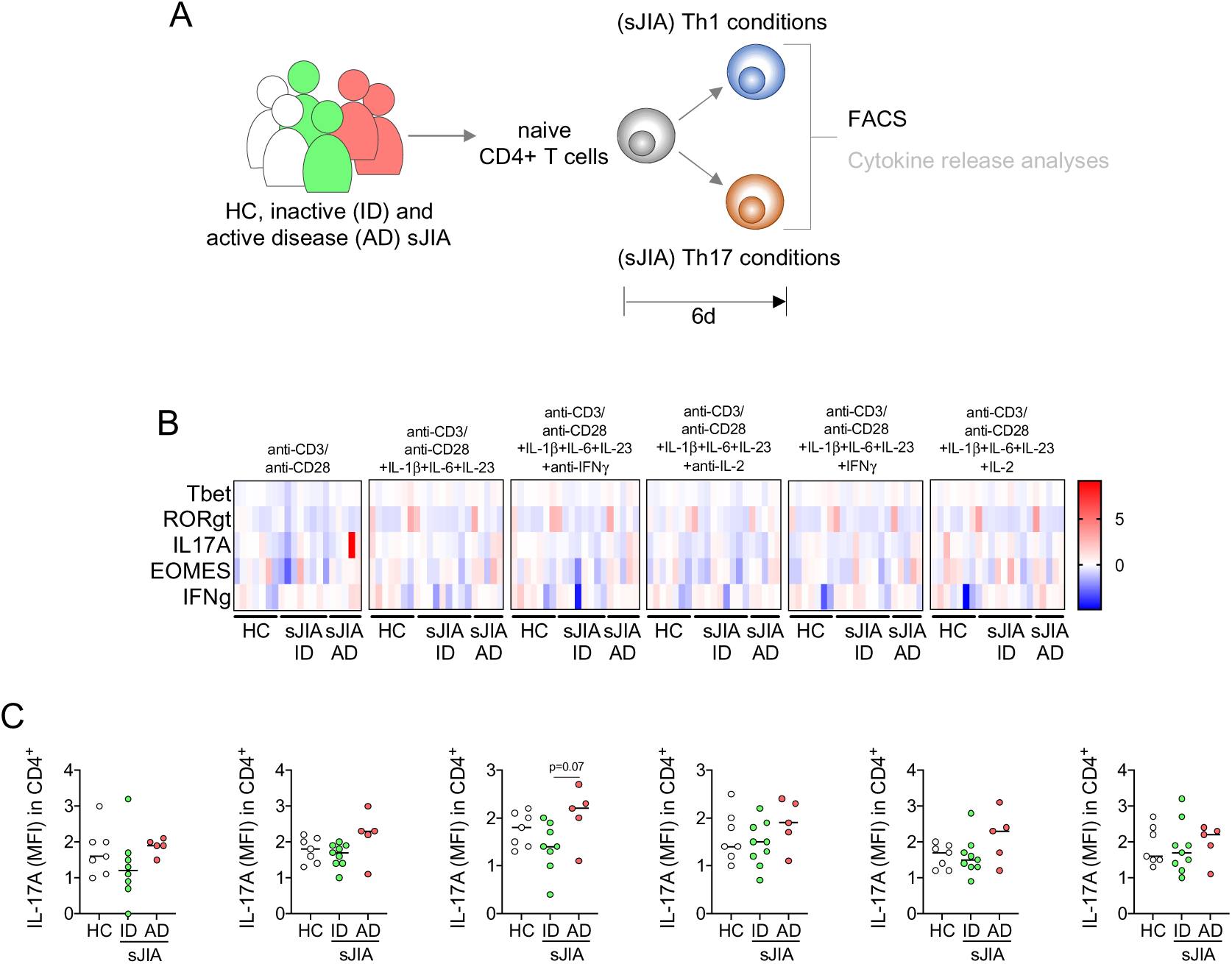
SJIA naïve CD4+ T helper (Th) cell differentiation is not skewed toward a T_h_17 fate or high IL-17A expression. **A**, Naive Th cells were isolated from pediatric healthy controls (HC, n=7), active disease (AD, n=5), and inactive disease (ID, n=8) sJIA patients (see table 1) and cultured under T0 (anti-CD3/anti-CD28) and different Th17 polarizing conditions. Following six days of culture and super-stimulation with phorbol 12-myristate 13- acetate/ionomycin (4h), cells were analyzed by flow cytometry. **B**, Heatmap of z-scores indicating T-bet, RORγt, IL-17A, IFNγ, and Eomesodermin (Eomes) expression (geometric mean fluorescence intensity (MFI) normalized to unstimulated samples) in cells arising from the indicated culture conditions and following super-stimulation as described. **C**, IL-17A expression (MFI) in cells as in **B** is shown. **B** and **C**, Each data point represents a value derived from an individual patient or HC. **C**, Lines in scattered dot plots indicate median values, data were analyzed by Kruskal-Wallis test followed by Dunn’s post hoc test.

**Figure S3.**
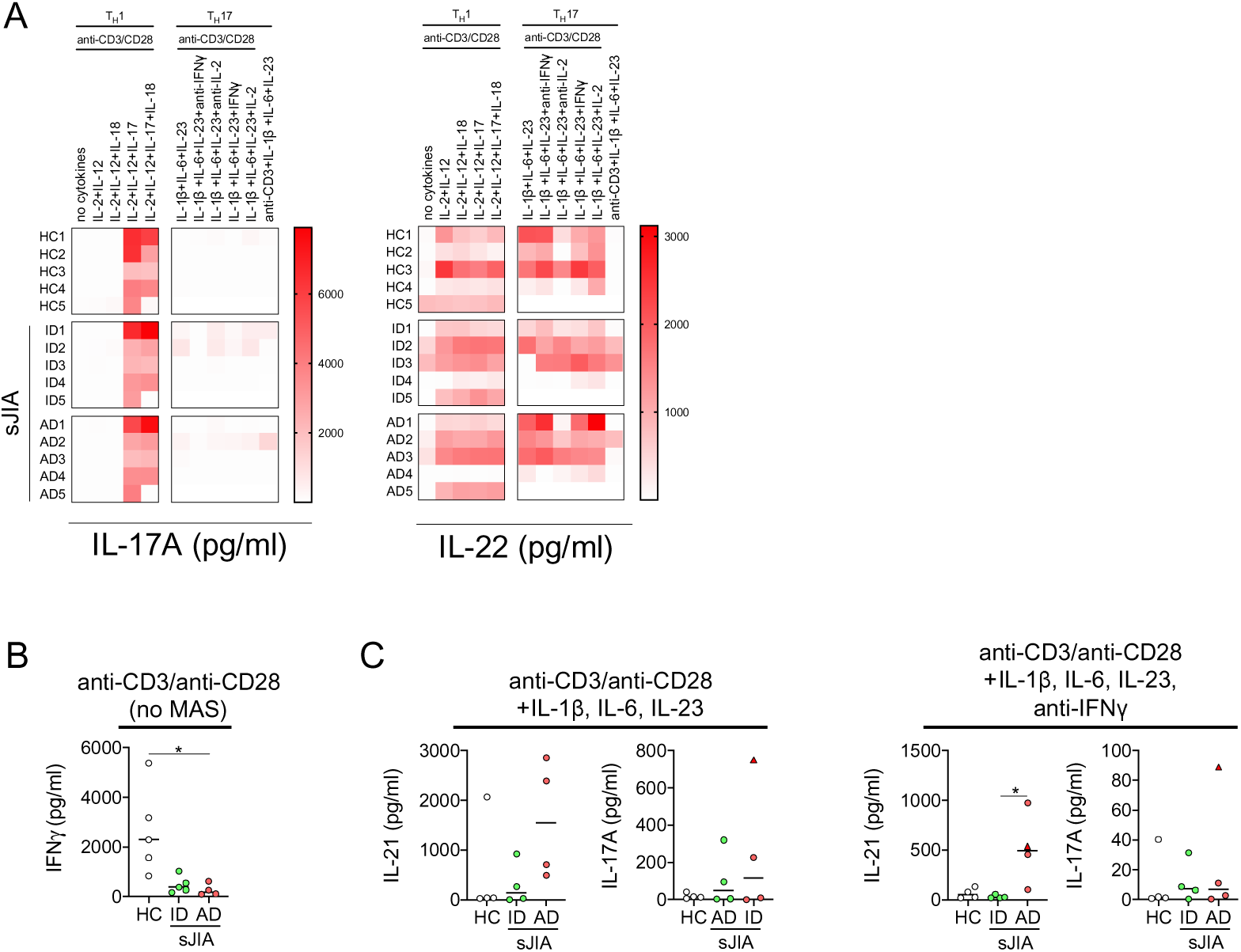
Developing CD4+ T helper type 1 (T_h_1) and type 17 (T_h_17) cells in systemic juvenile idiopathic arthritis (sJIA) do not release elevated levels of IL-17A or IL-22. **A**-**C**, Naive Th cells were isolated from pediatric healthy controls (HC, n=7), active disease (AD, n=5), and inactive disease (ID, n=8) sJIA patients (see table 1) and cultured under T0 (anti-CD3/anti-CD28) and different Th1 and Th17 polarizing conditions as indicated. On day six of culture, cytokine release (IFNγ, IL-17A, IL-21, IL-22) into culture supernatants following phorbol 12-myristate 13-acetate/ionomycin super-stimulation was analyzed by multiplexed bead array assay. **A**, Heatmaps of IL-17A (left panel) and IL-22 (right panel) release (pg/ml) by naive CD4+ cells following culture under the indicated T0, Th1, and Th17 polarizing conditions and super-stimulation as described. **B**, IFNγ release following Th0 polarization and super-stimulation with one patient developing MAS subsequent to the analyses excluded from the data set. **C**, IL-21 (left panels) and IL-17A (right panels) release by naive CD4+ cells cultured under the indicated Th17 polarizing conditions and following super-stimulation. Data is shown in scatter dot plots of the measured cytokine concentrations in pg/ml. **A**-**C**, Each data point presents a value derived from an individual patient or HC. **B** and **C,** Lines indicate median values. Data were analyzed by Kruskal-Wallis followed by Dunn’s post hoc test; * = *P* < 0.05.

**Figure S4.**
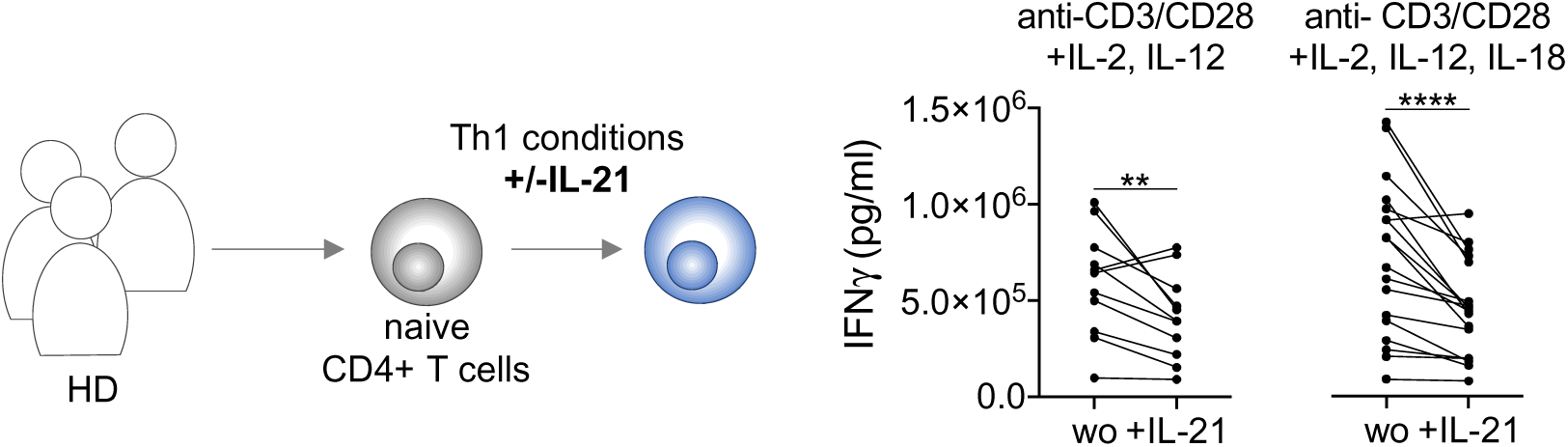
IL-21 restricts IFNγ release by developing CD4+ T helper type 1 (Th1) cells derived from naïve adult healthy donor CD4+ T cells. Naïve Th cells were isolated from adult healthy donors (HD, n=18) and cultured under Th1 polarizing conditions in the absence or presence of recombinant human IL-21. IL-18 was included to somewhat mimic the sJIA-cytokine environment. On day six of culture, IFNγ release into culture supernatants following phorbol 12-myristate 13-acetate/ionomycin super-stimulation was analyzed by ELISA. Scatter dot plots show IFNγ release in pg/ml by naive HC CD4+ cells cultured under the indicated Th1 polarizing conditions (left: n=11, right: n=18) in the absence (wo) and presence (+IL-21) of IL-21 and following super-stimulation as described. Each data point presents a value derived from an individual HD and data were analyzed by Wilcoxon signed-rank test; ** = *P* < 0.01, **** = *P*<0.0001.

**Figure S5.**
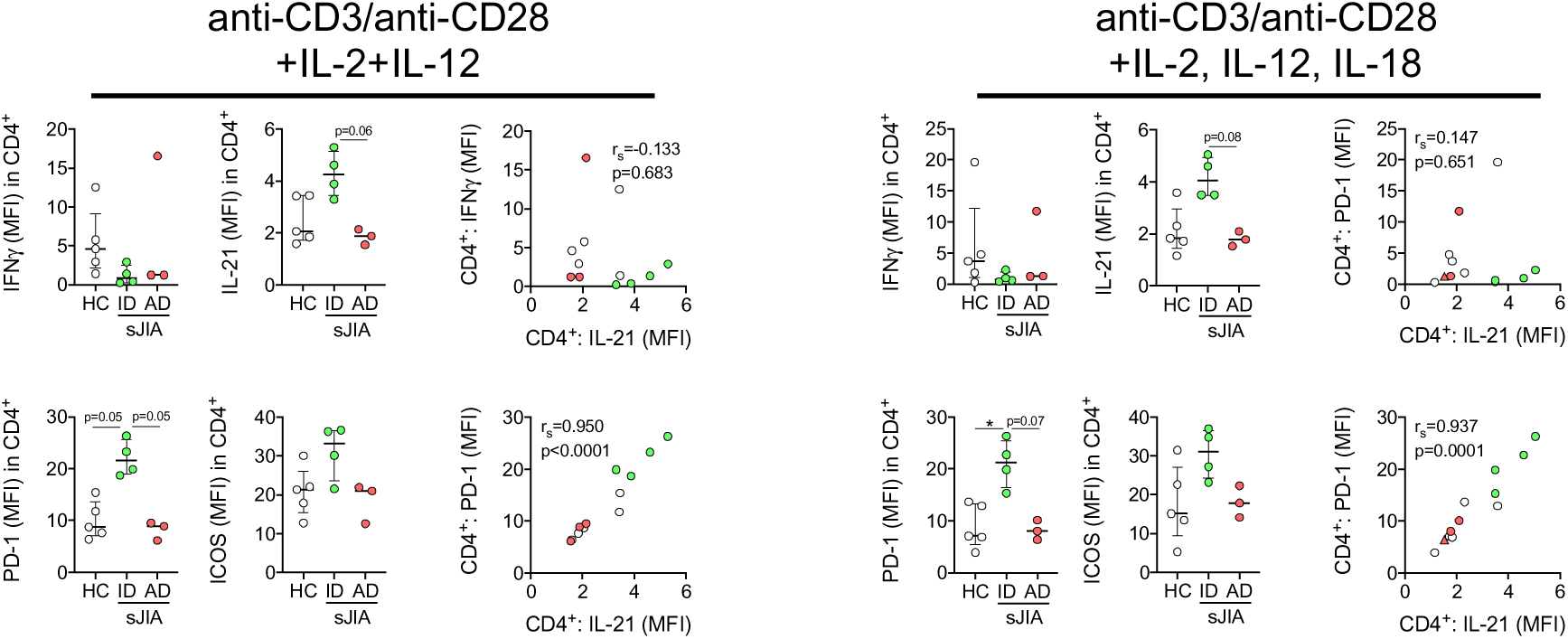
Early CD4+ T helper (Th) cell differentiation in systemic juvenile idiopathic arthritis (sJIA) is shifted toward a (PD-1+ICOS+CXCR5-) peripheral T helper (Tph) cell phenotype. Naïve Th cells were isolated from pediatric healthy controls (HC, n=5), active disease (AD, n=3), and inactive disease (ID, n=4) sJIA patients (see table 1) were cultured under T0, Th1, and T follicular helper (Tfh) / peripheral T helper (Tf/ph) polarizing conditions. Following six days of culture and super-stimulation with phorbol 12-myristate 13- acetate/ionomycin (4h), cells were analyzed by flow cytometry. IFNγ, IL-21, PD-1, and ICOS expression (MFI, left and middle columns) and correlation of IL-21 and IFNγ or PD-1 expression (MFI, right columns) by naïve CD4+ T cells cultured under the indicated conditions and following super-stimulation. Each data point presents a value derived from an individual patient or HC. Lines in scatter dot plots indicate median values. Data in left and middle columns were analyzed by Kruskal-Wallis followed by Dunn’s post hoc test; * = *P* < 0.05. Correlation data were analyzed by simple linear regression (OLS) and F-test; r_s_ (r-squared value) and *P*-values are indicated.

**Figure S6.**
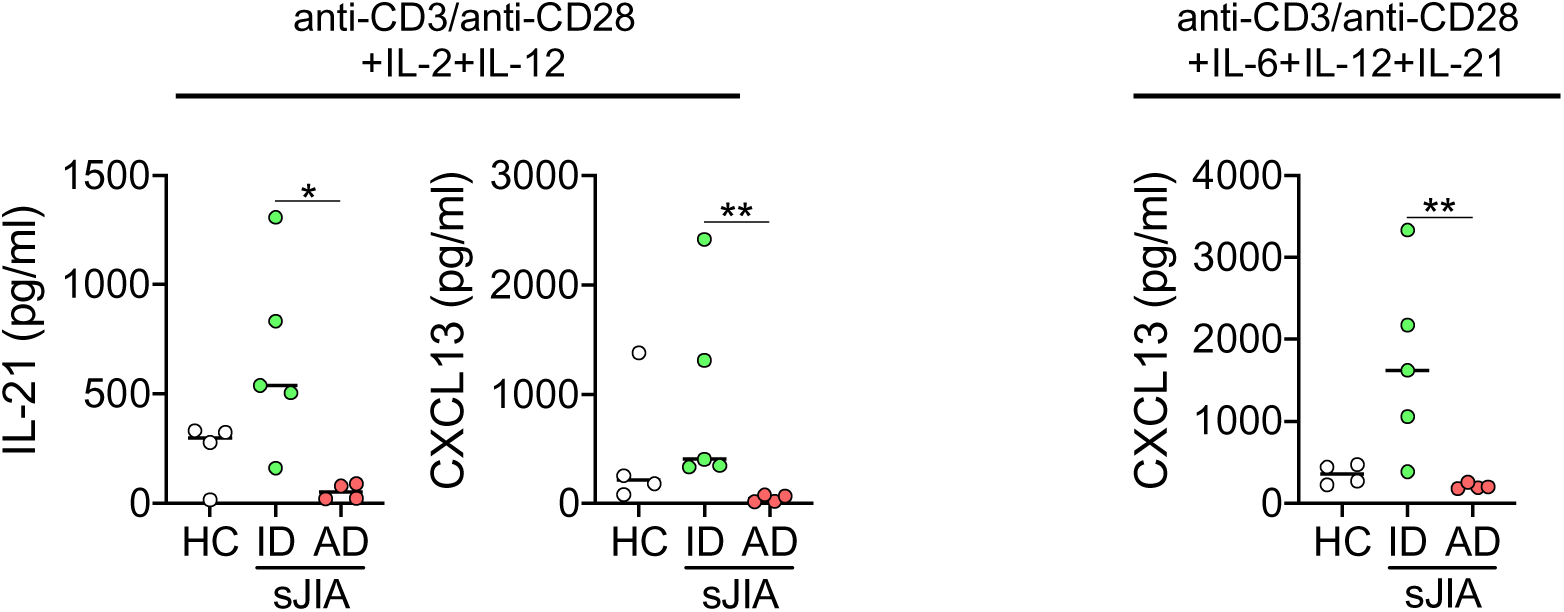
Developing CD4+ T helper type 1 (Th1) cells in systemic juvenile idiopathic arthritis (sJIA) release elevated levels of IL-21 and CXCL13. Supernatants of naïve Th cells isolated from pediatric healthy controls (HC, n=4), active disease (AD, n=4), and inactive disease (ID, n=5) sJIA patients (see table 1) cultured under indicated Th1 and T follicular helper (Tfh) / peripheral T helper (Tf/ph) polarizing conditions (6d) and super-stimulation with phorbol 12-myristate 13-acetate/ionomycin (4h) were analyzed by multiplexed bead array assay for IL-21 and CXCL13 expression. Data is shown in scatter dot plots of the measured cytokine expression in pg/ml. Each data point presents a value derived from an individual patient or HC. Lines indicate median values and data were analyzed by Kruskal-Wallis test followed by Dunn’s post hoc test; * = *P* < 0.05, ** = *P*<0.01.

**Figure S7.**
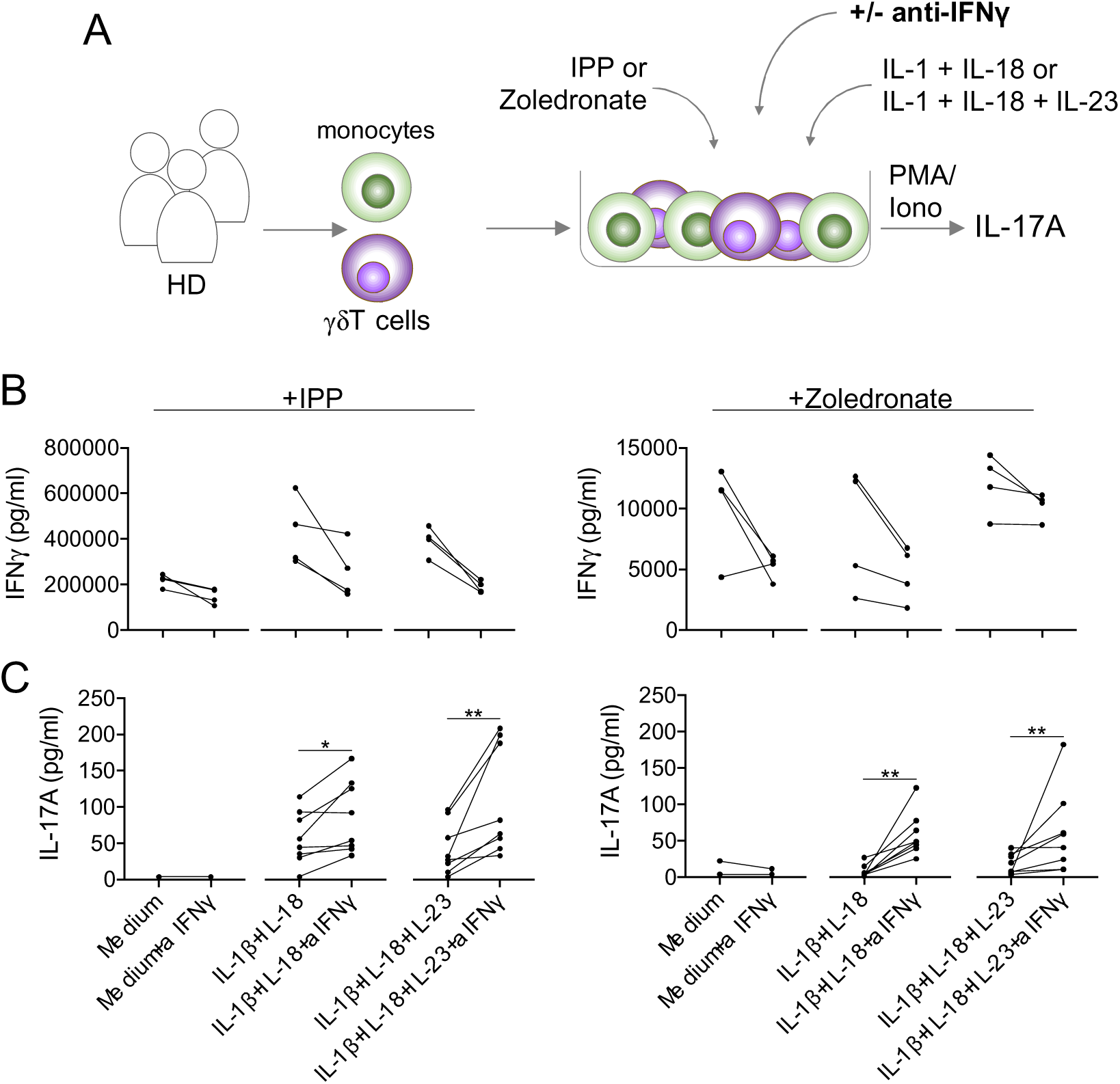
Low IFNγ benefits γδ T cellular IL-17A release. **A**-**C**, γδT cells and monocytes were isolated from adult healthy donors (HD) (n=4-8) and co-cultured in indicated conditions supplemented with isopentenyl pyrophosphate (IPP) or zoledronate and in the absence or presence (+aIFNγ) of an IFNγ-neutralizing antibody for five days. IFNγ-depletion (**B**) and paralleling IL-17A release (**C**) following super-stimulation with phorbol 12-myristate 13- acetate/ionomycin. was assessed by ELISA. Each data point presents a value derived from an individual HD and data were analyzed by Wilcoxon signed-rank test; * = *P*<0.05, ** = *P*<0.01.

